# Implementation of Uniform Processes of Care Across an Academic Health System During the Initial COVID-19 Pandemic Surge and Their Association with Outcomes

**DOI:** 10.1101/2023.06.01.23290807

**Authors:** JM Siner, C Price, M Villanueva, C Dela Cruz, S Honiden, J Johnson, CJ Winterbottom, MJ Franco, J Topal, D Mcmanus, M Malinis, L Tanoue, F Lorusso, M Phadke, F Li, S Sureshanand, K Churchwell, M Holmes, TJ Balcezak, G Desir

**Affiliations:** Yale University School of Medicine; Yale New Haven Hospital; Yale New Haven Health; Bridgeport Hospital; Greenwich Hospital

## Abstract

**Introduction:** Disparities in care delivery and outcomes are common in healthcare in the United States. The SARS-CoV-2 pandemic in the spring of 2020 in the United States and around the world resulted in a surge in the need for acute and critical care services for patient with acute respiratory disease. Many individual hospitals and health systems were unprepared for this surge of patients with a novel and acute respiratory disease which may have exacerbated pre-existing disparities. To prepare for this challenge the Yale New Haven Health System developed a response to the SARS-CoV-2 pandemic in 2020 which was multifactorial including: 1) Implementation of a uniform COVID management protocol across the care continuum, 2) Precise criteria for hospital and Intensive Care Unit (ICU) and Stepdown Unit (SDU) admission, 3) Augmented ICU and SDU bed availability, 4) Implemented load balancing across the entire health system. To understand the impact of these interventions we reviewed and compared mortality across the Yale-New Haven Health System both between hospitals and to national data. We also analyzed administration of medications to understand local adherence to the COVID-19 management protocol implemented during the initial wave of the pandemic.

**Methods:** This investigation is an observational, retrospective study of 3,112 patients infected with SARS-CoV-2 during the first wave of the pandemic in southern Connecticut and Rhode Island. All COVID-19 admissions to the Yale New Have Health System from March through June of 2020 were included. Patients all received care at a hospital within the Yale New Haven Health System which has 2693 beds across 7 campuses in southern Connecticut and Rhode Island. The primary outcome was in-hospital mortality for patients with COVID-19. Demographics were extracted as well as specific data associated with process of care including timing of administration of Tocilizumab, aspirin, and corticosteroids. Transfers between hospitals within the health system were identified. Mortality rates were compared between the central tertiary care hospital and the smaller community and community teaching hospitals using logistic regression to adjust for patient factors.

**Results:** Analysis of process of care metrics including time to Tocilizumab, aspirin, and corticosteroids shows adherence of recommended processes of care across Yale New Haven Health System. The overall mortality rate of 15.9% was lower than published national comparators. Hospital mortality rates compared between the central tertiary care center and smaller hospitals within the system were similar when adjusted for multiple patient factors including race and ethnicity.

**Conclusions:** In this investigation of COVID-19 outcomes in an academic health system with geographic and social diversity, we find that the observed low mortality rate was consistent across the health system. We propose that this is in part related to consistency of care and structural factors such as load balancing. We believe that these findings highlight the potential value of implementing uniform processes designed to reduce noise and bias in clinical judgment.

## Introduction

The scope of the SARS-CoV-2 pandemic presented a unique challenge for healthcare delivery characterized by a novel infectious agent and an immune naïve population. The initial major outbreaks in Lombardy (Italy) and New York City (USA) were characterized by rapid inundation of hospitals with a large number of patients with COVID-19 which presented primarily as pneumonia and hypoxemic respiratory failure due to Acute Respiratory Distress Syndrome (ARDS). ^1^ Initial reports of high hospital mortality were concerning particularly for mechanically ventilated patients.^2^ The modern healthcare system in the United States has limited experience with addressing urgent acute care and critical care for a large number of patients as occurs during a pandemic surge. In addition, healthcare delivery in the United States is replete with disparities in care that have been well characterized across multiple different locations, populations and care settings and there was immediate concern that these underlying disparities could be exacerbated in the setting of surge in need ^3^.

We sought to evaluate the impact of a high level of standardization and integration and coordination of care across a healthcare system on the outcomes for all COVID-19 admissions during the initial surge. A prior analysis of the initial wave of the pandemic within this health system had demonstrated age-adjusted in-hospital mortality for discharged patients was not significantly different among racial and ethnic groups. ^4^ We sought to further understand, the variance in outcomes across Yale New Haven Health System and analyze specific factors which could help us understand the extent of integrated and standardized care.

Yale New Haven Health System (the “System”) has several factors which promoted standardization and coordination of care and outcomes across the hospitals within the system. Structurally YNHHS developed an incident command system that developed and published guidelines for care for all health professionals including physicians, nurses, and respiratory care (Table 4). There were specific guidelines for hospital admission from the Emergency Department (ED) and for admission to intensive care unit (ICU) and Stepdown Unit (SDU) level care specifically related to oxygen requirements. The guidelines also emphasized use of High Flow Nasal Cannula (HFNC) rather than non-invasive ventilation (Bipap), and adherence to *accepted* evidence-based standard care of critically ill patients with ARDS (low tidal volume ventilation, pronation therapy). Importantly, despite reports of a very high mortality for COVID-19, specifically in patients who required mechanical ventilation, the System critical care group elected to approach care limitation and end of life planning no differently than the pre-existing practice with ARDS. Structurally, the System cancelled elective surgery early, and markedly increased Medical ICU and Medical Step-Down Unit bed capacity across the health system and actively used this capacity to load balance ICU case volume between hospitals within the System. In addition to accepting internal transfers, several of the hospitals accepted transfers from southwestern Connecticut hospitals outside the health system that had capacity challenges.

## Methods

The study design is an observational, retrospective study of patients with SARS-CoV-2 who received care at a Yale New Haven Health System (YNHHS) Hospital and were admitted between March 1 and June 30 of 2020. Yale New Haven Health System is a five-hospital academic health system with 7 distinct campuses with a total of 2693 beds and includes a tertiary care hospital as well as community teaching hospitals and community hospitals serving a diverse patient population with facilities across southern Connecticut and western Rhode Island. The Yale University Institutional Review Board (IRB) approved this investigation. Patient were identified based on an index admission during the specific time period associated with the presence of SARS-CoV-2 on RT-PCR testing (SARS-CoV-2 testing was either from a test developed at Yale-New Haven Hospital or by a Connecticut Department of Public Health State laboratory for those admitted in the early in the surge). All SARS-CoV-2 admissions between March 1, 2020 and June 30, 2020 were included. Repeat admissions with SARS-CoV-2 positive PCR were excluded (all patients admitted to a YNHHS hospital receive COVID-19 testing upon admission during this time period). Transfers to inpatient hospice at the same hospital (which include a discharge and readmission to the same location within the same hospital), were combined with the index admission for data purposes into a single admission. All data was extracted from the Epic EMR. Demographics were extracted as well as length of stay and information to calculate the Charlson Comorbidity Index; the Rothman score was calculated on all patients and already present within the EMR. The Rothman score functions as severity of illness score and combines physiologic data, lab data and nursing assessments. Of note “race” and “ethnicity” within our EMR are separate questions and so there can be overlap in the two populations. Also extracted was data associated with process of care specifically timing of administration of Tocilizumab, corticosteroids, and aspirin relative to the time of index hospital admission. Transfers between hospitals within the System were identified. Transfers from other hospitals in the state outside the System (also for load balancing) were treated similar to de-novo admissions and not counted as transfers. The primary outcome was in-hospital mortality for patients with COVID-19. Length of stay was calculated and combined into a single outcome for those who were transferred from one hospital to another within the System. The cohort of patients that was transferred for either load balancing or care needs were analyzed with their mortality ascribed to the receiving hospital. The transfer group was also analyzed on its own to further understand the impact of load balancing. To specifically understand how outcomes varied across the System we performed a multivariable logistic regression with mortality as the outcome for the central tertiary care hospital compared to aggregated outcomes from the other hospitals in the System. The regression was adjusted for variables known to be associated with mortality (age, gender, race, ethnicity, Rothman score, Charlson index and mechanical ventilation use).

## Results

In total, 3112 patients with SARS-CoV2 were admitted to one of the System hospitals between March 1, 2020 and June 30, 2020. Among these 3112 admissions, 33 were transferred from one of the smaller hospitals (B, C, D, E) to the tertiary care referral center (A) for either advanced therapies or load balancing or both (Table 2). Age and other demographic characteristics varied across the hospitals. While the number of patients transferred was low compared to the overall number of admissions, they all were deemed to be at high-risk for mortality as demonstrated by the frequency of mechanical ventilation (30 of 33 transfers) (Table 2). For the entire cohort of COVID-19 patients, the mean age was 64.3 years old with frequent presence of the comorbidities, hypertension (70.8%), congestive heart failure (25.1%), diabetes (39.7%). The admitted population with COVID-19 identified their race as 48.4% white, 26.4% Black or African American. For ethnicity, the admitted population, with COVID-19 26.2%, identified as Hispanic or Latino.

The overall mortality in the System was 15.9% for this early surge and among those requiring mechanical ventilation it was 31.3%. To determine the implementation of our COVID-19 protocol we chose to analyze process of care metrics around administration of specific medications that were part of the COVID-19 protocol. The percentage of patients who received a protocol recommended medication during the first 7 days after hospital admission (Tocilizumab, aspirin, or corticosteroids) is shown in Table 1. Since we could not reliably identify when patients met criteria retrospectively, the numbers are presented as a percentage of the entire population. While there was variability among the hospitals with regards to the percentage of patients who received the Tocilizumab within 7 days across the system, a substantial proportion of the patients received Tocilizumab at all of the hospitals and a negligible number would have otherwise received it as it is not part of the standard pre-pandemic sepsis or ARDS guidelines. Aspirin was included in the COVID-19 Treatment Protocol at a later date based on evolving data and corticosteroids were the last to be included in the COVID-19 Treatment Protocol. Analysis of the transfer of patients from the smaller hospitals to the larger tertiary care center shows substantial patient movement either for loading balancing, CRRT(Continuous Renal Replacement Therapy), or ECMO (Extracorporeal Membrane Oxygenation (Table 2). Nearly all of the transferred patients were critically ill, as demonstrated by the near uniform presence of mechanical ventilation (91%) with an associated higher expected high mortality rate (Table 2). Overall mortality rates across the Yale-New Haven Health system was 15.9% of all admissions and mortality was uniform across the health system (Table 1). Specifically, in a logistic regression model that adjusted for age, Charlson index score, Rothman Score (initial), race, ethnicity, gender, and requirement for mechanical ventilation there was no difference in the mortality rates between the central tertiary care hospital and the smaller hospitals in the health system (Table 3).

**Tables 1.** Clinical Characteristics and Outcomes of All Patients:

| Characteristic | Health System<br>(n= 3112) | Hospital A<br>(n = 1669) | Hospital B<br>(n = 858) | Hospital C<br>(n= 461) | Hospitals D & E<br>(n= 124) |
| --- | --- | --- | --- | --- | --- |
| Pre-Pandemic ICU Capacity Medical (Total) |  | 57 (178) | 16 (36) | 11 (11) | 16 (30) |
| Age (yrs) (mean, sd) | 64.29 (18.53) | 64.83 (17.93) | 63.27 (19.11) | 62.92 (19.89) | 69.18 (15.99) |
| Female Sex (n, %) | 1559 (50.10%) | 863 (51.71%) | 438 (51.05%) | 202 (43.82%) | 56 (45.16%) |
| Race (n, %) |  |  |  |  |  |
| Black or African American | 821 (26.38%) | 531 (31.82%) | 241 (28.09%) | 35 ( 7.59%) | 14 (11.29%) |
| Other/Unknown | 786 (25.26%) | 260 (15.58%) | 289 (33.68%) | 213 (46.20%) | 24 (19.35%) |
| White or Caucasian | 1505 (48.36%) | 878 (52.61%) | 328 (38.23%) | 213 (46.20%) | 86 (69.35%) |
| Ethnicity: Hispanic or Latino (n, %) | 814 (26.16%) | 322 (19.29%) | 286 (33.33%) | 185 (40.13%) | 21 (16.94%) |
| Charlson Score (mean, sd) | 1.81 (2.28) | 2.04 (2.37) | 1.75 (2.20) | 1.11 (2.02) | 1.73 (2.15) |
| BMI (mean, sd) | 29.70 (8.31) | 29.93 (8.53) | 30.01 (8.60) | 28.37 (5.90) | 29.29 (10.21) |
| Hypertension (n, %) | 2202 (70.76%) | 1276 (76.45%) | 587 (68.41%) | 242 (52.49%) | 97 (78.23%) |
| Asthma (n, %) | 638 (20.50%) | 378 (22.65%) | 190 (22.14%) | 53 (11.50%) | 17 (13.71%) |
| CHF (n, %) | 783 (25.16%) | 475 (28.46%) | 201 (23.43%) | 68 (14.75%) | 39 (31.45%) |
| Diabetes (n, %) | 1236 (39.72%) | 708 (42.42%) | 343 (39.98%) | 133 (28.85%) | 52 (41.94%) |
| Initial Rothman Score (mean, sd) | 61.20 (24.26) | 59.23 (23.94) | 61.84 (24.35) | 70.46 (21.83) | 49.12 (26.02) |
| Mechanical Ventilation n(%(95% CI)) | 568<br>(18.3%<br>(16.9,19.6)) | 329<br>(19.7%<br>(17.8,21.6)) | 161<br>(18.8%<br>(16.2,21.4)) | 49<br>(10.6%<br>(7.8,13.4)) | 29<br>(23.4% (15.9,30.8)) |
| Tocilizumab within 7 days of Admission n(%(95% CI)) | 1218<br>(39.1%<br>(37.4,40.9)) | 845<br>(50.6%<br>(48.2,53)) | 222<br>(25.9%<br>(22.9,28.8)) | 116<br>(25.2%<br>(21.2,29.1)) | 35<br>(28.2% (20.3,36.1)) |
| Corticosteroids within 7 days of Admission n(%(95% CI)) | 270<br>(8.7% (7.7,9.7)) | 164<br>(9.8%<br>(8.4,11.3)) | 68<br>(7.9%<br>(6.1,9.7)) | 23<br>(5.0%<br>(3.0,7.0)) | 15<br>(12.1% (6.4,17.8)) |
| Aspirin within 7 days of Admission n(%(95% CI)) | 1014 (32.6%<br>(30.9,34.2)) | 617 (37%<br>(34.7,39.3)) | 246 (28.7%<br>(25.6,31.7)) | 102 (22.1%<br>(18.3,25.9)) | 49 (39.5%<br>(30.9,48.1)) |
| Length of Stay in Days (median, IQR) | 8.1<br>(4.3 – 15.3) | 9.2<br>(5.0 – 17.5) | 8.0<br>(4.1 – 15.2) | 5.8<br>(3.1 – 9.7) | 7.0<br>(4.1 – 13.3) |
| Transfers Received (n) | 33 | 33 | 0 | 0 | 0 |
| Transfers Sent (n) | 33 | 0 | 6 | 24 | 3 |
| Mortality n(%(95% CI)) | 494 (15.9%<br>(14.6,17.2)) | 272 (16.3%<br>(14.5,18.1)) | 139 (16.2%<br>(13.7,18.7)) | 52 (11.3%<br>(8.4,14.2)) | 31 (25% (17.4,32.6)) |
| Mortality among those Mechanically ventilated n(%(95% CI)) | 178 (31.3%<br>(27.5, 35.2)) | 93 (28.27%<br>(23.4, 33.1)) | 60 (37.27%<br>(29.8, 44.7)) | 16 (32.65%<br>(19.5, 45.8)) | 9 (31.03% (14.2, 47.9)) |

**Table 2.** Characteristics and Outcomes for Patients Who Underwent Inter-hospital Transfer:

| Characteristic | Transfers to Referral Center (A)<br>(n= 33) |
| --- | --- |
| Sending Hospital | D = 3, C= 24, B= 6, |
| Age (yrs) (mean, sd) | 52.70 (14.37) |
| Female Sex (n, %) | 8 (24.24%) |
| Race (n, %) |  |
| Black or African American | 4 (12.12%) |
| Other/Unknown | 20 (60.61%) |
| White or Caucasian | 9 (27.27%) |
| Ethnicity: Hispanic or Latino (n, %) | 21 (63.64%) |
| Charlson Index Score (mean, sd) | 0.53 (1.05) |
| BMI (mean, sd) | 30.57 (6.66) |
| Hypertension (n, %) | 17 (51.52%) |
| Asthma (n, %) | 2 (6.06%) |
| CHF (n, %) | 2 (6.06%) |
| Diabetes (n, %) | 10 (30.30%) |
| Rothman Score (mean, sd) | 60.66 (22.42) |
| Mechanical Ventilation (n, %) | 30 (90.91%) |
| Tocilizumab within 7 days (n, %) | 30 (90.91%) |
| Corticosteroids within 7 days (n, %) | 1 (3.03%) |
| Aspirin within 7 days (n, %) | 8 (24.24%) |
| Length of Stay in Days (median, IQR) | 34.3 (22.1 – 54.0) |
| Mortality (n, %) | 13 (39.39%) |
| Mortality among those Mechanically ventilated (n, %) | 11 (36.67%) |

**Table 3.** Logistic Regression of Mortality at Tertiary Care Hospital compared to Aggregated Data from the Remaining 4 System Hospitals Adjusted for Patient Factors.

| Characteristic | Odds Ratio | P-value | 95% Confidence Intervals |
| --- | --- | --- | --- |
| Other Hospitals vs Tertiary Hospital | 1.004 | 0.9762 | (0.796 – 1.265) |
| Age | 1.062 | <0.0001 | (1.051 – 1.073) |
| Rothman Index (initial) | 0.969 | <0.0001 | (0.964 – 0.974) |
| Charlson Index Score | 1.059 | 0.0162 | (1.011 – 1.019) |
| Gender (Female vs. Male) | 0.682 | 0.0014 | (0.540 – 0.863) |
| Race: Black or African-American vs White or Caucasian | 0.892 | 0.4395 | (0.667 – 1.192) |
| Race: Other or Unknown vs White or Caucasian | 1.320 | 0.2195 | (0.847 – 2.057) |
| Ethnicity: Hispanic or Latino vs Not Hispanic or Not Latino | 0.818 | 0.3711 | (0.527 – 1.270) |
| Mechanical Ventilation (Yes vs No) | 3.740 | <.0001 | (2.848 – 4.912) |

**Table 4:** Pandemic Preparation & Protocols

| System Preparation | Protocols & Guidelines |
| --- | --- |
| Development of COVID Treatment Team to Develop Guidelines | Medication Protocols |
| Early cancellation of Elective Surgery | Low tidal volume ventilation adherence |
| Conversion of cancer hospital to COVID-19 ICU | Standard approach to goals of care |
| Expansion of ECMO Capability | HFNC supported (over Bipap) and no early intubation |
| Expansion of Tele-ICU (including daytime coverage) |  |
| Expanded ICU Staffing model (physician and nursing) |  |

## Discussion

Reported outcomes for COVID-19 patients admitted to the hospital have been highly variable. In particular several early analyses show high mortality at the onset of the pandemic with improvements during the course of the first wave. ^5,6^ While disease specific care improved over time, patient volume and local disease incidence had a major impact on mortality with the simplest explanation being that hospital strain is persistent factor in outcomes. ^7^ What none of the studies have been able to address is whether structural or logistical *preparation* for or response to a surge impacts local outcomes (Table 4). However, one analysis suggested that pre-existing structural factors such as baseline ICU and hospital bed capacity may contribute to reduced mortality. ^8^

Early in the pandemic there were no disease specific therapies that clearly modified the morbidity or mortality of the disease. Without disease specific therapies the ability of a hospital to provide adequate supportive care would be expected to be the primary driver of outcomes. Therefore, structural issues such as quality and uniformity of care and ability to mitigate the impact of high volumes on clinical care would be expected to be substantial contributors to any observed variation in outcomes. In view of the consistency of mortality rates across the System, we considered whether some of the same factors including standardized care pathways, standardized admission criteria, and load balancing that appeared to protect against disparities might have contributed to improved outcomes.^4^

We hypothesized that, in the setting of healthcare system strain, adherence to standards of care and reduced variation could explain the lack of variation in outcomes according to race and ethnicity previously described for the System.^4^ Through similar mechanisms, this reduced variation in care could reduce decrease mortality and minimize variation among the System hospitals. Specifically, we hypothesized that standardization of care (indications and usage for Tocilizumab and steroids and aspirin), ICU/SDU admission criteria, along with system-wide load balancing and intensive preparation collectively would contribute to improved and comparable system-wide mortality. While is it clear that the implementation of the practice standards was not identical, the ability to achieve broadly similar practice with new medications in the setting of a highly disruptive pandemic does demonstrate a substantial standardization even though the medication administration rates are not identical. In this current investigation of outcomes in an academic health system with geographic and patient diversity we describe consistency of delivered care across the health system (process) combined with structural load balancing and augmented capacity as potential contributors to comparatively low mortality rates that were consistent across the health system (Table 3). The logistic regression analysis adjusting for patient factors demonstrated that the outcomes (mortality rate) were no different despite variations in severity of illness and race and ethnicity among other differences.

Studies from the initial surge show that nationally there were high mortality rates in the early surge of 2020.^5,9^ Multiple investigators have reviewed the relationship between caseload and outcomes and some have suggested that hospital based resources related to ICU bed availability may be an important component of mortality when adjusted for local COVID19 incidence.^6,8^ An investigation by Block et al stratified by caseload and ICU availability and found that mortality ranged from 15.2% in the lowest quintile to 22.7% for the highest quintile of patient volume and a similar study found 21.4% mortality in the initial surge.^9,10^ Using Blocks’s methodology, the System would be expected to have a hospital mortality consistent with that highest (top) quintile for mortality (22.7%), which is higher than the observed rate of 15.9% for the patients admitted in our cohort.^10^ A separate investigation suggested that structural factors may be less important, and a primary driver of mortality may be the community infection rate.^6^ This certainly may be correct given the lack of disease specific therapies but we cannot explain low mortality rates as due to a low community infection rate as 3 of the 5 hospitals in the System were in areas with substantial surges and had very high community transmission in the early pandemic although not all the System hospitals are in the same community. Nearly all of the System hospitals experienced high community levels of spread which has been associated with worse mortality, and yet despite *baseline* variations in practice and ICU bed availability the outcomes were nonetheless similar. A key element in many of the other investigations were that the hospitals subject to the early wave of the pandemic were concentrated in the Northeast and were at highest risk for poor outcomes. Overall, the mortality rate at the hospitals in the System showed remarkable consistency with regards to mortality despite variation in COVID-19 volume as assessed by COVID19 admission per bed (Block 2021)(Table 3).^10^ While many factors impacted outcomes, investigations from multiple different hospitals and systems of outcomes in the initial wave generally demonstrated a mortality rate in excess of 20%, whereas the mortality rate in our cohort was substantially lower.^2,10-13^

The standardization of admission criteria and the use and timing of medications across the health system reflects a joint approach and ability to communicate and disseminate practice standards. Taken together these system approaches functioned as a form of proactive preventative care undertaken to avoid the risk of inundation of the hospitals above their capacity to provide the usual standard of care which was as important, or more important, than the medical interventions at that time (Table 4). The System has both academic and community teaching hospitals that collectively participate in setting system standards. It would be difficult to determine which of the multiple factors resulted in the observed homogeneity of outcomes but certainly the process of care measures were similar (given the rapidly evolving practices during the pandemic) and the outcomes were similar across the system (Table 3) suggesting that the combination of process of care standardization with load balancing was able to equalize outcomes despite varying case burdens (Table 3). Importantly, the System hospitals provide a substantial portion and often the predominance of healthcare provided in many of their communities. As such, expanding capacity and providing standardized care was direct service to their communities likely contributed to a reduction in disparities. We believe these findings are an important contribution because it has been conjectured that standardization and load balancing locally and regionally might have improved the ability of the US healthcare system to respond to a pandemic and we believe that these data from our experience show that this approach potentially does have merit if it can be executed in a standardized and coordinated and inclusive fashion.^14^

### Potential Limitations

While a large health system the plurality of the admissions were at the central tertiary care facility which would bias any analysis towards structural factors at the main hospital. It remains possible that additional factors related to viral inoculum and local infection rates and likelihood and timing of presentation and admission acuity patients to the hospitals in the health system could have impacted hospitalization rates as well as mortality. The interplay and balance of the impact of logistics (ICU/SDU bed availability), load balancing, versus care standardization) is not possible to delineate as these interventions were collectively a bundle. However, we do believe that all these factors undertaken in a prospective and preventative fashion due to concerns about the resources being overwhelmed served to standardize care and collectively contributed to the observed uniform good outcomes.

## Data Availability

All data produced in the present work are contained in the manuscript.

## Acknowledgements

The authors would like to thank the following for their assistance in preparing the analyzing the data and manuscript: James Dziura MPH PhD, Margaret Pisani MD, MPH. Importantly the authors also would like to thank <u>all</u> those from YNHHS including both the front line staff, administration and leadership who supported them for their contributions to the pandemic response all of whom worked together to expand our capacities and support each other and our patients.

## Literature Cited

1. Distante C, Piscitelli P, Miani A. Covid-19 Outbreak Progression in Italian Regions: Approaching the Peak by the End of March in Northern Italy and First Week of April in Southern Italy. Int J Environ Res Public Health 2020;17(9). DOI: 10.3390/ijerph17093025.

2. Richardson S, Hirsch JS, Narasimhan M, et al. Presenting Characteristics, Comorbidities, and Outcomes Among 5700 Patients Hospitalized With COVID-19 in the New York City Area. JAMA 2020;323(20):2052–2059. DOI: 10.1001/jama.2020.6775.

3. Arul K, Mesfin A. The Top 100 Cited Papers in Health Care Disparities: a Bibliometric Analysis. J Racial Ethn Health Disparities 2017;4(5):854–865. DOI: 10.1007/s40615-016-0288-y.

4. McPadden J, Warner F, Young HP, et al. Clinical characteristics and outcomes for 7,995 patients with SARS-CoV-2 infection. PLoS One 2021;16(3):e0243291. DOI: 10.1371/journal.pone.0243291.

5. Horwitz LI, Jones SA, Cerfolio RJ, et al. Trends in COVID-19 Risk-Adjusted Mortality Rates. J Hosp Med 2021;16(2):90–92. DOI: 10.12788/jhm.3552.

6. Asch DA, Sheils NE, Islam MN, et al. Variation in US Hospital Mortality Rates for Patients Admitted With COVID-19 During the First 6 Months of the Pandemic. JAMA Intern Med 2021;181(4):471–478. DOI: 10.1001/jamainternmed.2020.8193.

7. Kadri SS, Sun J, Lawandi A, et al. Association Between Caseload Surge and COVID-19 Survival in 558 U.S. Hospitals, March to August 2020. Ann Intern Med 2021;174(9):1240–1251. DOI: 10.7326/M21-1213.

8. Janke AT, Mei H, Rothenberg C, Becher RD, Lin Z, Venkatesh AK. Analysis of Hospital Resource Availability and COVID-19 Mortality Across the United States. J Hosp Med 2021;16(4):211–214. DOI: 10.12788/jhm.3539.

9. Fried MW, Crawford JM, Mospan AR, et al. Patient Characteristics and Outcomes of 11 721 Patients With Coronavirus Disease 2019 (COVID-19) Hospitalized Across the United States. Clin Infect Dis 2021;72(10):e558–e565. DOI: 10.1093/cid/ciaa1268.

10. Block BL, Martin TM, Boscardin WJ, et al. Variation in COVID-19 Mortality Across 117 US Hospitals in High- and Low-Burden Settings. J Hosp Med 2021;16(4):215–218. DOI: 10.12788/jhm.3612.

11. Argenziano MG, Bruce SL, Slater CL, et al. Characterization and clinical course of 1000 patients with coronavirus disease 2019 in New York: retrospective case series. BMJ 2020;369:m1996. DOI: 10.1136/bmj.m1996.

12. Cummings MJ, Baldwin MR, Abrams D, et al. Epidemiology, clinical course, and outcomes of critically ill adults with COVID-19 in New York City: a prospective cohort study. Lancet 2020;395(10239):1763–1770. DOI: 10.1016/S0140-6736(20)31189-2.

13. Suleyman G, Fadel RA, Malette KM, et al. Clinical Characteristics and Morbidity Associated With Coronavirus Disease 2019 in a Series of Patients in Metropolitan Detroit. JAMA Netw Open 2020;3(6):e2012270. DOI: 10.1001/jamanetworkopen.2020.12270.

14. Mitchell SH, Rigler J, Baum K. Regional Transfer Coordination and Hospital Load Balancing During COVID-19 Surges. JAMA Health Forum 2022;3(2):e215048. DOI: 10.1001/jamahealthforum.2021.5048.

